# Phased amplicon multiplex sequencing for cost-effective detection of high-risk human papillomavirus from cervical samples

**DOI:** 10.64898/2026.07.27.26359029

**Authors:** Dipesh Solanky, Charlotte Low, Christine L. Hathaway, Stephen Cherne, Elizabeth Brown, Thesla Palanee-Phillips, Ruanne V. Barnabas, Roby P. Bhattacharyya, Brittany M. Berdy, Jonathan Livny

**Affiliations:** Broad Institute of Massachusetts Institute of Technology and Harvard University, Cambridge, MA, USA; Division of Infectious Diseases, Mass General Brigham, Boston, MA, USA; Department of Medicine, Harvard Medical School, Boston, MA, USA; University of Washington School of Medicine, Seattle, WA, USA; Department of Pathology, University of Washington, Seattle, WA, USA; University of Washington School of Public Health, Seattle, WA, USA

## Abstract

Access to accurate cost-effective technologies for typing high-risk human papillomaviruses (hrHPV) is critical to expand cervical cancer screening and inform vaccination strategies. Compared with clinical-standard quantitative polymerase chain reaction (qPCR) assays, HPV genotyping by next-generation sequencing (NGS) provides greater flexibility, scalability, and genotype specificity. We have developed a method for HPV genotyping, HPV Phased Amplicon Multiplex Sequencing (PhAM-Seq), that uses combinatorial barcoding of amplicons with short, variable-length inline sequences to enable higher throughput and lower per-sample costs than conventional amplicon sequencing approaches. We evaluated HPV PhAM-Seq using degenerate and type-specific primers targeting the L1 and E6–E7 gene loci in a blinded cohort of 170 cervical samples previously typed by the Seegene Anyplex II HPV28 Detection qPCR assay. Across eight common hrHPV types (HPV16, 18, 31, 33, 35, 45, 52, and 58), HPV PhAM-Seq demonstrated >80% overall agreement with qPCR using degenerate L1-targeting primers, with the highest sensitivity for HPV16, 31, 33, and 58. Sensitivity for HPV35, 45, and 52 improved to ≥85% with type-specific primers targeting genes E6/E7. Parallel processing and sequencing enables a single technician to assay hundreds of samples per week at a reagent cost of around $10 USD per sample, with laboratory automation and sequencing on higher-output platforms enabling further scaling and cost reduction to a scale amenable to population-level surveillance. We include a detailed SOP; tools for primer design, sequencing library construction, and sample tracking; and all scripts needed for data analysis to ensure HPV PhAM-Seq can be readily implemented for scalable, cost-effective hrHPV genotyping or extended to other similar applications.

## Introduction

Infection with oncogenic, or high-risk (hr), genotypes of human papillomavirus (HPV) is associated with up to 99.7% of cases of cervical cancer, the fourth leading cancer in women by both incidence and mortality worldwide [1]. HPV is a double-stranded DNA virus with an 8000 base-pair (bp) genome that encodes 6 early-expressed proteins (E1, E2, E4, E5, E6, and E7) and 2 late proteins (L1 and L2) that form the viral capsid [2]. Carcinogenesis with hr genotypes of HPV infection is largely driven by viral integration into the host genome, leading to transcription of oncogenic proteins E6 and E7 [3]. HR genotypes HPV16 and HPV18 cause 70% of HPV-related cervical cancer. These, combined with 6 additional types (HPV31/33/35/45/52/58), cause approximately 96% of HPV-related cervical cancers, with a greater proportion of HPV35 in Africa (3.6%) compared to other regions (0.6–1.6%) [4].

The current approaches to screen for HPV-related cervical cancer and precancer include visual inspection with acetic acid, cervical cytology (i.e., Papanicolaou testing), and nucleic acid-based hrHPV testing. Globally, visual inspection with acetic acid continues to be the most recommended approach in resource-limited settings [5]. However, adding nucleic acid-based hrHPV testing to cervical cytology increases its sensitivity for detection of cervical precancer to *>*90% compared to 50–79% for cytology alone [6, 7]. Additionally, randomized controlled trial data from multiple high-income industrial countries (HICs) show that the incorporation of hrHPV testing to cervical cancer screening corresponds to decreases in cervical precancer and invasive cervical cancer incidence [8–12]. The World Health Organization (WHO) has called for the elimination of cervical cancer globally, recommending that 70% of women aged 35 and older undergo cervical cancer screening with a high-quality HPV test at least twice in their lifetime [13]. The WHO also recommends that hrHPV testing for this purpose prioritizes restricted genotyping (RG) of the 8 hr types (HPV16, 18, 31, 33, 35, 45, 52, and 58) most commonly linked to cervical cancer [14]. The prevalence and distribution of oncogenic HPV types can vary widely across communities within a population [15]. Understanding this variability through hrHPV genotyping of populations is critical to optimizing equitable distribution of resources for cervical cancer screening, HPV testing, and treatment of HPV-related cervical cancer and precancer.

Reliable hrHPV testing on a population level can also be leveraged to guide global HPV vaccination efforts. Vaccination against HPV, including bi- and tetravalent formulations, has been shown to reduce the risk of acquisition of oncogenic HPV types in young adults by 80–90% [16–18], and there are ongoing efforts to increase HPV vaccine availability and uptake in multiple low- and middle-income countries (LMICs) [19]. Although 7 of the 8 hr types in the RG panel are targets in all current HPV vaccine formulations (with the exception of HPV35), inequities in vaccine availability and coverage limit the potential impact of vaccination in reducing HPV prevalence among young adults worldwide [20]. Within this current reality, cost-effective, sustainable HPV testing and typing at scale can mitigate the adverse impact of limited vaccine supply by identifying communities with the greatest burden of hrHPV to prioritize for vaccine allocation while concurrent efforts are taken to augment vaccine resources. Additionally, as HPV vaccination programs increase, accurate HPV genotyping of populations pre- and post-vaccination is key to assessing vaccine efficacy, duration of protection, emergence of novel variants, and type replacement [21].

Despite the crucial role of hrHPV testing in reducing the cervical cancer burden—both as a screening tool for cervical disease as well as informing vaccination program strategy—several barriers continue to limit its use in low- and middle-income countries (LMICs), including in regions with the highest cervical cancer rates, including sub-Saharan Africa [2, 22]. Among these are the high costs of commercial typing assays that can be deployed in LMICs, which can vary from $5-71 per sample, as well as the additional instrument costs of up to $150,000 and/or specialized training required for both commercial and non-commercial methods [23, 24]. Moreover, the ability to multiplex samples using these approaches is often very limited, making them impractical in large-scale HPV typing efforts, such as those measuring infection dynamics or vaccine efficacy in large populations.

Several groups have recently developed methods for HPV genotyping using next-generation sequencing (NGS) of polymerase chain reaction (PCR)-amplified DNA, or amplicons [25–29]. Amplicon NGS offers several advantages over PCR-based HPV genotyping. First, the direct sequence information generated by NGS enables greater analytical specificity through sequence-level confirmation, improved sensitivity through reduction of false-negative results, and the ability to detect genotypes and variants beyond pre-designed primer targets [30–32]. Second, amplicon sequencing offers substantially greater primer flexibility, permitting inclusion of a larger number of primers within a single PCR reaction than is feasible with real-time PCR [33], and enabling the use of barcoding strategies to further increase target multiplexing capacity [34]. Although these NGS-based methods demonstrated high sensitivity for hrHPV detection and strong concordance with established PCR-based diagnostic assays, each presents critical barriers that have hindered their broad implementation. These barriers include per-sample costs often exceeding those of commercial assays (Table 1 [25–27, 35, 36]); limited sample size; restricted capacity for multiplexing samples during processing and/or sequencing; lengthy and operationally complex laboratory workflows; and the absence of readily accessible, method-compatible tools for primer design and downstream data analysis.

**Table 1.** Per-sample costs of selected next-generation sequencing (NGS)-based HPV genotyping platforms.

| Study [25–27, 35, 36] | Platform / Method | Throughput Context | Cost per Sample (USD) |
| --- | --- | --- | --- |
| Barzon et al. (2011) | 454 pyrosequencing | 164 samples/run | \$55.20* |
| Wagner et al. (2019) | Illumina MiSeq | 96 samples/run | \$13.10 |
| Wagner et al. (2019) | Ion Torrent | 768 samples/run | \$6.00 |
| Chan et al. (2020) | Oxford Nanopore sequencing | 201 samples/run | \$50.77 |
| Lippert et al. (2021) | Ion Torrent | 60 samples/run | \$88.76† |
| Jitvaropas et al. (2023) | Illumina MiSeq | 38 samples/run | \$50.00 |

\* €40 converted using the 2011 exchange rate (approximately 1 EUR = 1.38 USD).
† €75.22 converted using the 2021 exchange rate (approximately 1 EUR = 1.18 USD).

To address the limitations of these approaches, we have developed HPV phased amplicon multiplex sequencing (HPV PhAM-Seq), a streamlined, scalable method that enables highly-multiplexed, cost-effective HPV genotyping by NGS. In this assay, regions of HPV genomes are amplified using primers that incorporate short DNA barcodes of variable lengths, after which barcoded amplicons are pooled and converted into sequencing libraries using indexed Illumina adapters. This combinatorial barcoding strategy substantially reduces cost and increases throughput by enabling hundreds of samples to be processed in parallel and sequenced simultaneously. As a result—even for small cohorts comprising dozens to hundreds of samples—the per-sample cost of HPV PhAM-Seq is a fraction of that of established PCR-based assays, and laboratory automation and sequencing on high-density platforms can further increase throughput and lower per-sample cost, offering scalability potentially compatible with population-level screening.

Here, we describe the development of HPV PhAM-Seq, including results from initial optimization experiments, performance evaluation in a blinded cohort of human cervical samples, and systematic troubleshooting to refine assay robustness. We present HPV PhAM-Seq as a streamlined, customizable, and scalable workflow designed for straightforward implementation in diverse laboratory settings. We include Google Workspace tools for primer design, sample tracking, library preparation management, and automated linkage of sample metadata to sequencing outputs, as well as all bioinformatics tools for data processing, genotype assignment, and reporting. Together, these resources enable end-to-end implementation of the platform, from primer design to library generation to data analysis.

## Materials and methods

### Primer design

Primers used for the first PCR of HPV PhAM-Seq encoded an 18-nucleotide Illumina TruSeq sequencing primer site (forward or reverse) at the 5’ end and a region targeting one or more HPV genotypes at the 3’ end (Fig 1). These 3’ sequences included degenerate MY11/09 primer sequences previously designed to target multiple genotypes [32], and type-specific primers targeting HPV18, HPV35, HPV45, or HPV52, designed using Geneious Prime [37] and Primer3 [38]. All HPV-specific binding regions within the primer were designed with an estimated melting temperature of 55°C. For some primers 1-2 additional nucleotides were added as inline barcodes between the Illumina and HPV sequences as shown in Fig 1. Sequences for these primers are found in S1 Table. A workbook for modifying these primers or generating new ones is available as described in the Data Availability Statement. Primers for the second PCR encode forward and reverse (P5 and P7) Illumina adapter and variable index sequences to facilitate sequencing of amplicons on an Illumina flow cell (Fig 1). Sequences for these primers are found in S2 Table.

**Figure 1.**
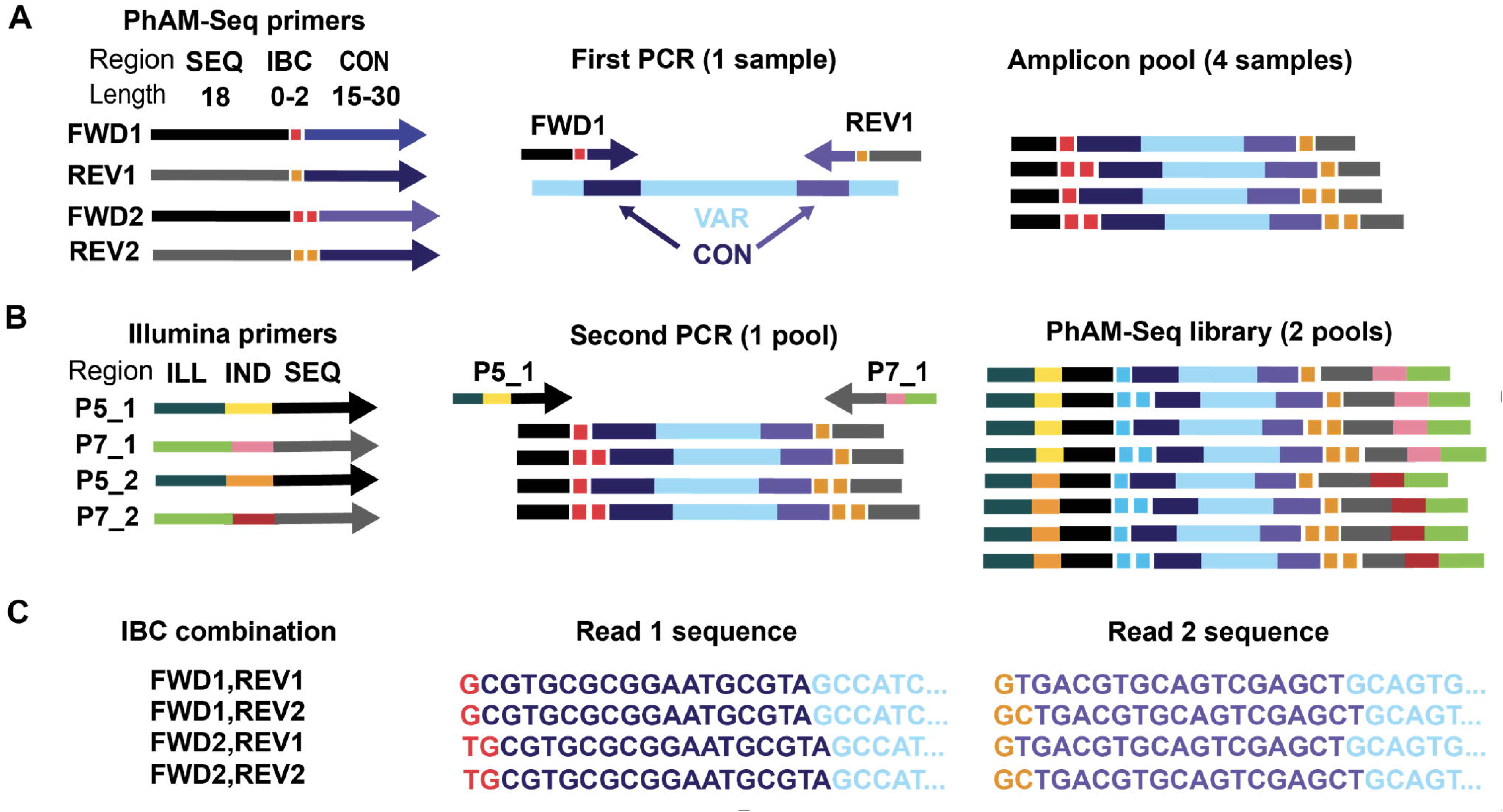
Schematic of PhAM-Seq sample barcoding and demultiplexing. A: PhAM-Seq primers target conserved sequences (CON) flanking a region of sequence variability (VAR). Each PhAM-Seq primer contains (i) an inline barcode (IBC) and (ii) a truncated binding site for one of the two TruSeq Illumina sequencing primers (SEQ). Amplicons for each sample are generated with one pair of PhAM-Seq primers. B: Amplicons with unique combinations of forward and reverse IBC are pooled and converted to Illumina sequencing libraries using primers targeting the SEQ regions that encode full-length P5 and P7 Illumina adapters (ILL) and index (IND) sequences. C: After sequencing, pools are demultiplexed by their Illumina index sequences, and individual samples are demultiplexed by their associated pairs of IBCs, with the length of each IBC inferred from the position of the CON sequences within each read.

### DNA isolation from cervical swabs

We evaluated the genotyping performance of the HPV amplicon sequencing platform using clinician-collected cervical samples collected in the Microbicide Trials Network(MTN)-020/ASPIRE Study, a phase 3, randomized, double-blind, placebo-controlled trial using a monthly vaginal ring containing dapivirine to prevent HIV-1 infection in sexually active women in Malawi, South Africa, Uganda, and Zimbabwe [39]. As part of the trial protocol, healthcare providers collected endocervical swabs from 2600 participants. Dry swabs stored at minus 80°C were shipped to the Pathology Laboratory at Harborview Medical Center. Swabs were suspended in 500 µL of phosphate-buffered saline (PBS), 200 µL of which was withdrawn for DNA extraction. DNA extraction was performed on the MagNA Pure 96 DNA and Viral NA Small Volume Kit (Roche, Basel, Switzerland). Extracted DNA samples were subsequently tested for the presence of low- and high-risk HPV types using a commercial semi-quantitative multiplex PCR assay (Anyplex II HPV28 Detection, Seegene, Seoul, South Korea), which can detect 28 HPV types and will be referred to as SG for the remainder of the manuscript. Authors did not have access to information that could identify individual participants during or after data collection.

### Assay controls

We obtained purified plasmids containing the entire genome of HPV16 to serve as a positive template control (American Type Culture Collection, Manassas, USA). To evaluate the performance of the assay on samples containing human DNA but negative for HPV DNA based on the reference assay, we also included samples that tested negative for any HPV type by the Seegene assay, referred to as “HPV-negative” controls. We included the positive template control and at least one sample of nuclease-free water as a no-template control in all experiments.

### PCR reactions

Between July 27 and August 2, 2025, 5 µL of stored purified DNA from each sample was combined with 20 µL of PCR master mix (25 µL total reaction volume) containing 5 µL of 5X Platinum II PCR Buffer (Thermo Fisher Scientific, Waltham, MA, USA), 0.5 µL of 10 mM dNTP mix (Thermo Fisher Scientific), 1 µL each of 5 µM forward and reverse HPV PhAM-Seq primers (S1 Table), 0.2 µL of Platinum II Taq Hot-Start DNA Polymerase (Thermo Fisher Scientific), and 12.3 µL of nuclease-free water. PCR cycling conditions were: 94°C for 2 min; 40 cycles of 94°C for 30 s, 55°C for 30 s, and 68°C for 1 min; followed by a final extension at 68°C for 5 min. Amplicons were purified using solid-phase reversible immobilization (SPRI) at 0.6× with AMPure XP beads (Beckman Coulter, Brea, CA, USA) and quantified using the Quant-iT™ dsDNA Assay Kit (High Sensitivity, Thermo Fisher Scientific). A subset ( 10–20%) of samples underwent size determination using a D1000 ScreenTape Assay on a 4200 TapeStation System (Agilent Technologies, Santa Clara, CA, USA). Samples were pooled according to in-line staggered or phased barcode combinations incorporated into both forward and reverse primers (0, 1, or 2 additional bases on each primer; 3 × 3 = 9 unique combinations). Prior to pooling, amplicons were normalized to equal input by mass to ensure equivalent contribution from each sample.

### Sequencing library generation, pooling, and sequencing

For each sample pool, 5 µL of pooled amplicons was combined with 18 µL of PCR master mix (23 µL total reaction volume) containing 2.5 µL of 10× AccuPrime PCR Buffer I (Thermo Fisher Scientific, Waltham, MA, USA), 0.2 µL of AccuPrime HiFi Taq DNA Polymerase (5 U/µL; Thermo Fisher Scientific), 1 µL each of 12.5 µM indexed Illumina P5 and P7 primers (Supplementary Table 2), and 15.3 µL of nuclease-free water. A 10-cycle second-round PCR (indexing PCR) was performed to append barcoded Illumina adapters. Cycling conditions were identical to those used in the first enrichment PCR, with the exception of 10 cycles instead of 40. Following amplification, indexed pools were purified using 0.6× SPRI with AMPure XP beads and quantified using the D1000 or HS D1000 ScreenTape Assay on a 4200 TapeStation System (Agilent Technologies, Santa Clara, CA, USA). Pools were normalized, combined at equivalent mass per pool to generate the final sequencing library, and subjected to a final 0.6× SPRI cleanup and TapeStation quality assessment prior to sequencing on an Illumina iSeq 100 System to yield 150-base, paired-end reads.

### Sequencing data processing and analysis

A curated HPV reference database was constructed by retrieving all annotated protein coding sequences from Papillomaviridae genomes in NCBI GenBank between 1.5Kb-9Kb in length and extracting L1, E6, and E7 genes using custom parsing scripts and labeled by HPV genotype. Sequencing reads were processed using Python-based scripts and standard bioinformatics tools, including Burrows–Wheeler Aligner (BWA) [40]. The complete pipeline—including all scripts, reference files, documentation and workbooks for sample tracking—is available as described in the Data Availability Statement. Reads were aligned to our curated HPV reference database of non-redundant L1, E6, and E7 gene sequences. Per-sample HPV types were summarized by counting reads assigned to each type and a sample was associated with an HPV type(s) if the sample reached or exceeded thresholds specified on the command line for 1) the total reads assigned to the sample (–min-total-reads), 2) the percentage of total read pairs assigned to the HPV genotype in the highest abundance (–min-pct-total), and 3) the percentage of all HPV read pairs assigned to the most abundant HPV genotype (–min-pct-typed). For each sample, up to 3 HPV types were reported, along with the percentage of reads assigned to each type. Additional information on generation of the HPV reference sequence database and sequence data processing and HPV genotyping is described in the S1 Appendix.

### Comparison of HPV genotyping results between assays

After running the pipeline on our sample cohorts, we determined agreement between HPV PhAM-Seq (HS) and SG for each sample, including HPV-negative samples. For each HPV type and HPV-negative samples as defined by SG, we grouped and tallied all concordant and discordant cases. Positive concordant cases (i.e., HAS+/SG+) were defined as detection of the same type in a sample by both assays or, for HPV-negative samples, absence of HPV DNA by both assays. Conversely, negative concordant results (i.e., HS-/SG-) were defined as absence of a specific HPV type in both assays; for HPV-negative samples, this corresponded to some HPV DNA—but not necessarily the same HPV type—detected by both methods. Discordant cases were defined as HS+/SG- or HS-/SG+ when there was disagreement between HS and SG. Positive percent agreement (sensitivity) was calculated as the proportion of HS-positive samples that were also SG-positive (HS+SG+ divided by the sum of HS+/SG+ and HS-SG+). Negative percent agreement (specificity) was calculated as the proportion of HS-negative samples that were also SG-negative ( HS-/SG-divided by the sum of HS-/SG- and HS+/SG-). Overall agreement was calculated as the proportion of concordant results among all samples (sum of HS+/SG+ and HS-/SG-samples divided by total sample number).

## Results

### PhAM-Seq Summary

Fig 1 and Fig 2 summarize the PhAM-Seq barcoding scheme and HPV PhAM-Seq assay workflow. Briefly, amplicons for each sample are generated by PCR using PhAM-Seq primers that contain a phased, inline barcode (IBC). Samples labeled with unique forward and reverse IBC combinations are pooled and converted to sequencing libraries using primers carrying Illumina flowcell adapter and index sequences. After sequencing, pools are first demultiplexed by their Illumina index, then individual samples are demultiplexed by their associated IBCs. Following IBC demultiplexing, all primer-derived sequences are trimmed, leaving only the amplified region for downstream analysis.

**Figure 2.**
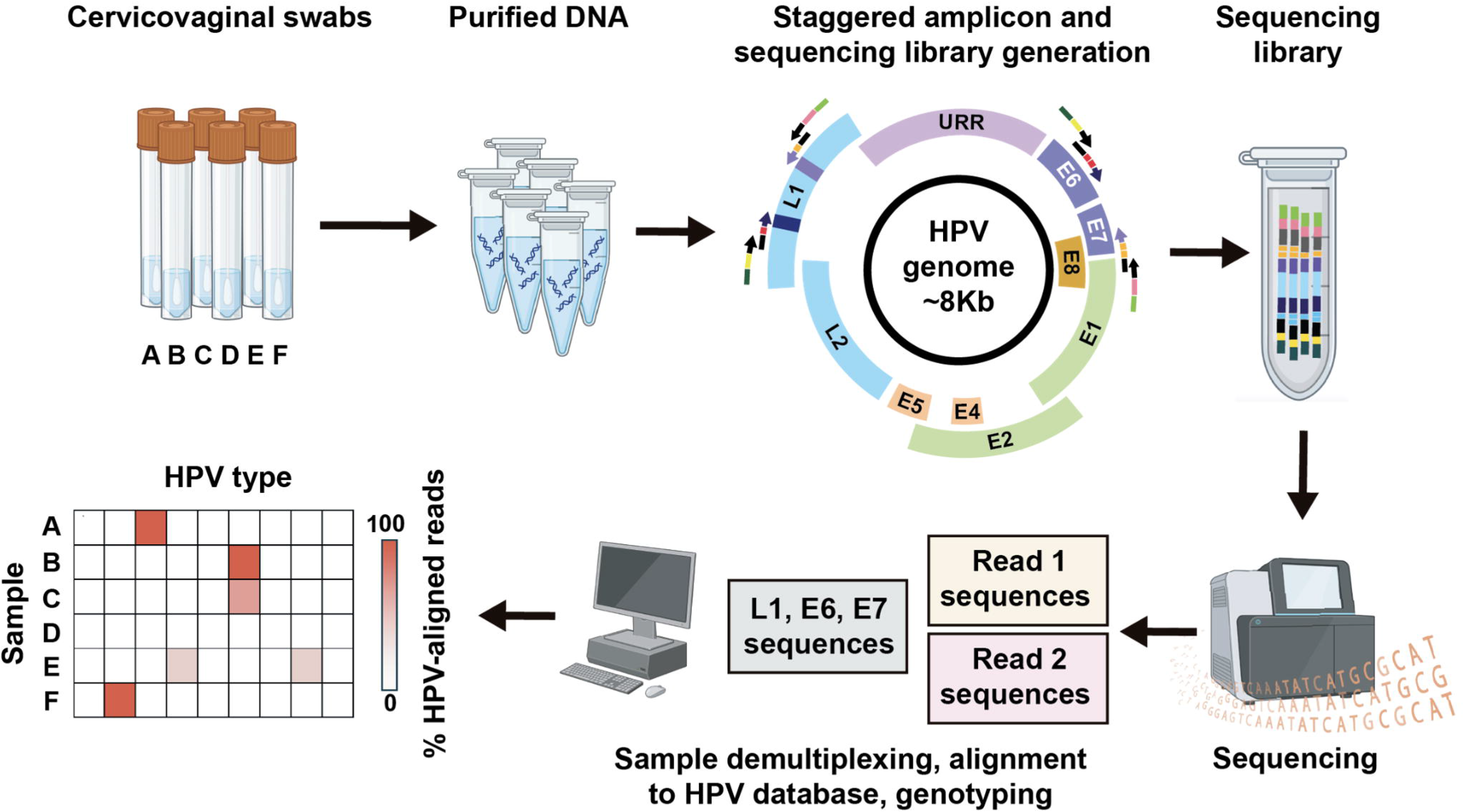
HPV PhAM-Seq workflow. Cervical swabs resuspended in a neutral buffer undergo DNA extraction and purification, after which a sequencing library is prepared through two sequential PCRs. All samples undergo a separate first enrichment. Samples are subsequently sorted into pools based on inline barcodes from the first enrichment and undergo a second enrichment to add unique Illumina indexes coupled with uniform adapter sequences to enable sequencing on an Illumina flow cell. Amplicons from all samples are pooled into a single, final sequencing library and loaded into an Illumina flowcell for sequencing. Sequencing reads are then demultiplexed by Illumina index-inline barcode combination, primer-derived sequences are removed, and the remaining sequences are aligned to an HPV sequence database and used to assign each sample to one or more HPV types.

Short, variable-length inline barcodes in PhAM-Seq offer three key advantages. First, combinatorial barcoding dramatically increases multiplexing complexity, such that the total number of uniquely identifiable samples scales multiplicatively with the number of forward IBCs, reverse IBCs, and unique Illumina indexes. Second, shorter primers reduce primer-primer interactions and off-target hybridization, improving amplification efficiency and fidelity. Third, staggered inline barcodes increase the base diversity of sequencing reads in each cycle, improving sequencing quality for low-diversity amplicon libraries. Additional information on the theoretical basis and scaling potential of IBCs is found in S1 Appendix.

### Validation of HPV PhAM-Seq on a blinded cohort

After validating and optimizing the workflow for HPV PhAM-Seq using plasmid-encoded HPV genomes, we tested the assay on a cohort of unique MTN-020/ASPIRE samples previously typed as mono-infections or HPV-negative by SG. We conducted the assay in a partially blinded fashion, wherein one investigator (BMB) selected 8-27 samples called by SG as one of each of the RG types, and 20 HPV-negative samples. A second investigator (DS) processed the samples and analyzed the results, without knowledge of the SG-assigned type for each sample by SG. In total, 170 cervical samples were processed but only samples above a minimum concentration after the first enrichment were pooled and sequenced.

Prior to the second enrichment, samples were divided into two parallel pooling strategies: one in which samples were normalized following the first enrichment prior to pooling, and another in which equal volumes from each sample were pooled without regard to sample concentration (i.e., non-normalized). Pools were sequenced and data was processed as described in the methods.

### Pipeline Threshold Determination

To determine the optimal parameters for analysis of HPV PhAM-Seq data, we compared type assignments across our cohort after running the pipeline iteratively varying 4 thresholds: 1) minimum total read depth, 2) maximum number of read pairs sampled for typing analysis, 3) minimum percent of total reads assigned to any HPV type (–min pct total), and 4) minimum percent of reads assigned to all HPV types that are assigned to a single type (–min pct typed). Modulating these thresholds impacted percent agreement with SG for each of the 8 RG types. We found that percent agreement with SG was maximized for all types when thresholds were set at 2500, 500, 1–2%, and 2%, respectively (Fig 3). We determined optimal thresholds by maximizing concordance with SG while minimizing spurious type calls arising from low read counts, which may reflect low-level background (e.g., minor amplicon carryover between samples).

**Figure 3.**
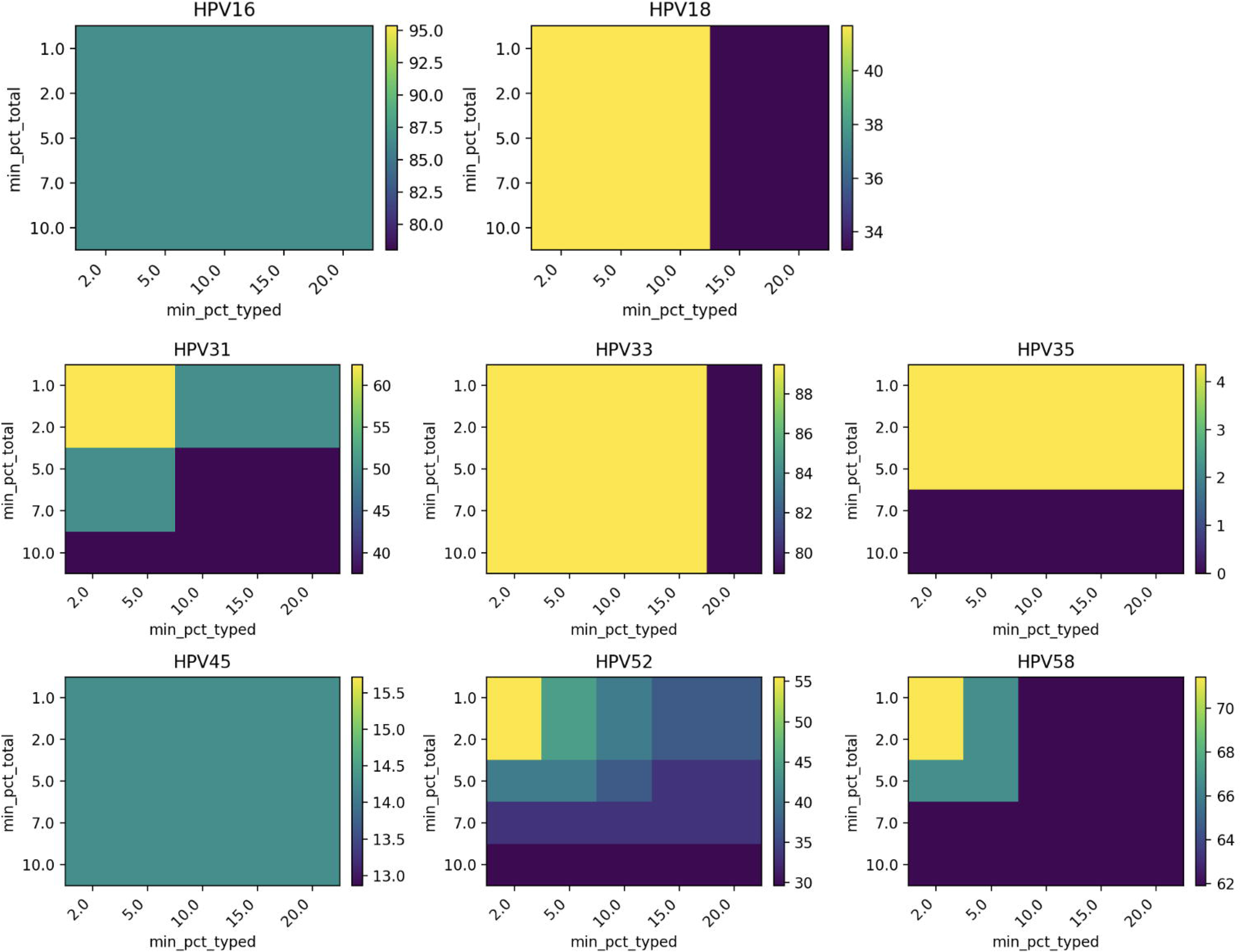
Read threshold optimization. Variation in percent concordance with SG for a given HPV type (color scale) with different thresholds for min-pct-total and min-pct-typed are shown. The pipeline was run at a minimum total read depth of 500 read pairs and maximum number of read pairs sampled for typing analysis of 2500 read pairs. HPV=human papillomavirus; min-pct-total = the percentage of total read pairs assigned to HPV genotype in highest abundance; min-pct-typed = the percentage of all read pairs assigned to any HPV genotype assigned to the HPV genotype in highest abundance.

### HPV PhAM-Seq on blinded samples demonstrates high overall agreement and type-dependent sensitivity

We evaluated concordance between HPV PhAM-Seq and SG using the optimized analysis parameters described above (Table 2). Negative and overall percent agreement with SG was *>*90% for all HPV types. Positive percent agreement with SG was more variable, ranging from *≥* 70% for HPV16/33/58 to *≤* 15% for HPV35/45 (Table 2).

**Table 2.**
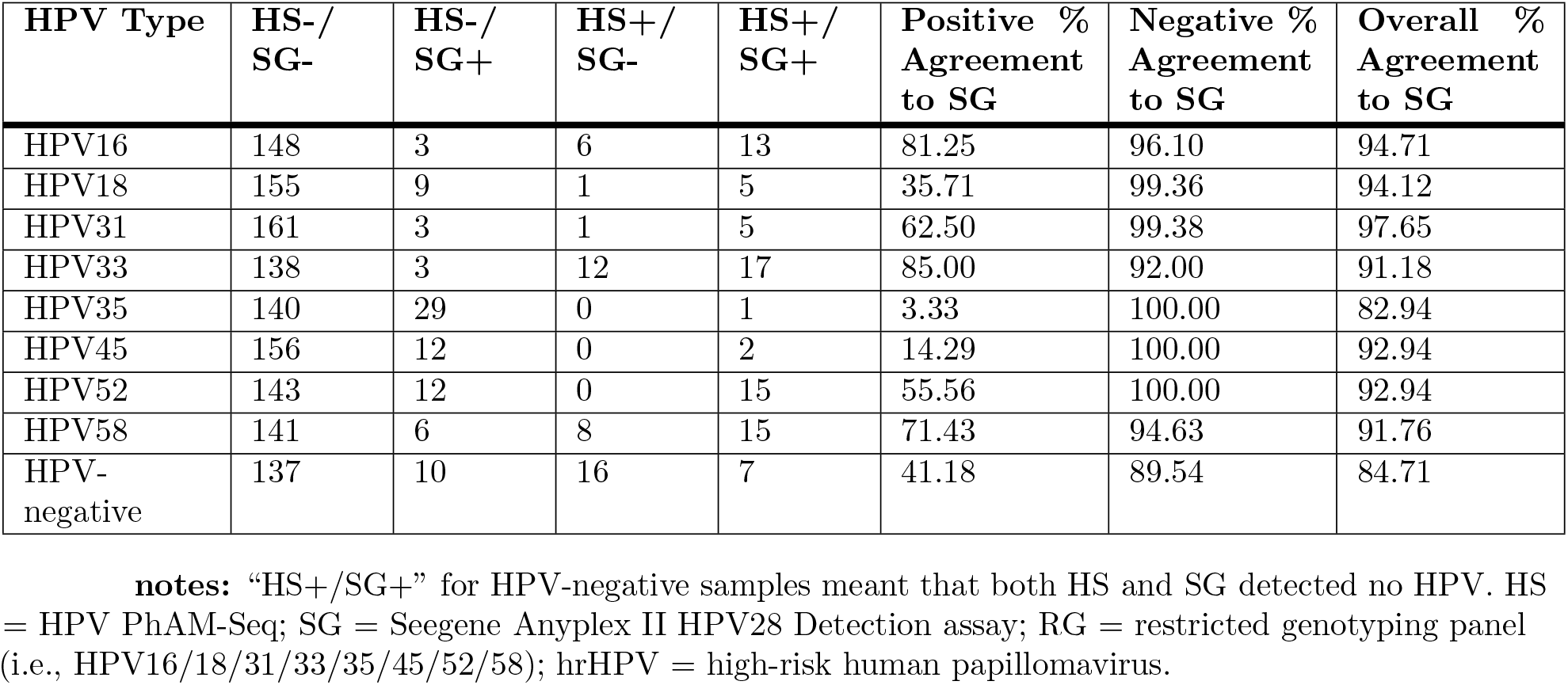
Agreement of HS with SG, stratified by the 8 RG hrHPV genotypes and HPV-negative.

Among the 30 SG-positive HPV35 samples, HPV PhAM-Seq identified HPV35 in one sample. In the remaining samples, HPV PhAM-Seq most commonly detected no HPV (n=13) or HPV71 (n=7). In one-third of SG+ HPV35 samples (n=10), a hr genotype other than HPV35 was identified by PhAM-Seq, including 5 samples that detected a high- or intermediate-risk type outside the SG panel (i.e., HPV53/70).

Among the 14 SG-typed HPV45 samples, HPV PhAM-Seq most commonly detected HPV72 (n=8) or HPV70 (n=4). Further, HPV PhAM-Seq detected HPV DNA in 10 samples reported as HPV-negative by SG, including hr types in 5 of these samples (HPV16/18/33/58, data not shown). In one sample classified as HPV18 by SG, HPV PhAM-Seq identified HPV85, an additional high-risk type not present on the SG panel. A summary of all discordant findings is shown in S1 Fig.

### Overall performance for hrHPV detection

When considering detection of any hrHPV type, including types outside the SG panel, HPV PhAM-Seq identified an hrHPV type in 95 (63%) of the 150 samples classified as hrHPV-positive by SG (Table 3). Of the 55 hrHPV samples identified by SG but not by HPV PhAM-Seq, 11 (20%) were excluded from sequencing due to insufficient DNA concentration after the first enrichment, and 68.2% (n=30) of the remaining 44 samples were typed by SG as HPV35, HPV45, or HPV52 (data not shown), consistent with the lower positive percent agreement observed for these types (Table 3). HPV PhAM-Seq detected hrHPV in five samples classified as HPV-negative by SG (data not shown).

**Table 3.** Summary of HS performance on detecting any hrHPV type relative to SG, including those outside the RG panel, comparing consideration of all HPV types detected versus only the top match.

| HPV Type | HS-/SG- | HS-/SG+ | HS+/SG- | HS+/SG+ | Positive % Agreement to SG | Negative % Agreement to SG | Overall % Agreement to SG |
| --- | --- | --- | --- | --- | --- | --- | --- |
| Any High-Risk Type Identified | 15 | 55 | 5 | 95 | 63.33 | 75.00 | 64.71 |

### Type-specific primers targeting L1 outside the MY region and E6–7 loci improves HPV PhAM-Seq detection of samples typed as HPV18/35/45/52 by SG

In addition to SG-typed HPV35/45 samples, HPV PhAM-Seq also demonstrated *<*65% positive agreement for SG-typed HPV18/31/52 samples (Table 2). We hypothesized that the relatively lower rate of detection for these types was potentially due to insufficient complementarity with the MY11/09 primers and/or L1 gene loss from viral integration into human chromosomal DNA. We therefore designed custom, type-specific primers targeting either other regions of the L1 gene or E6-7 gene regions for each of these types. The results are summarized in Table 4. We tested 29 of the original 30 SG-typed HPV35 samples assayed in the blinded validation. HPV35-specific primers targeting the L1 gene outside the MY region identified HPV35 in 25 of 29 samples (86.2%), compared with a positive percent agreement of 3.3% using MY11/09 primers.

**Table 4.**
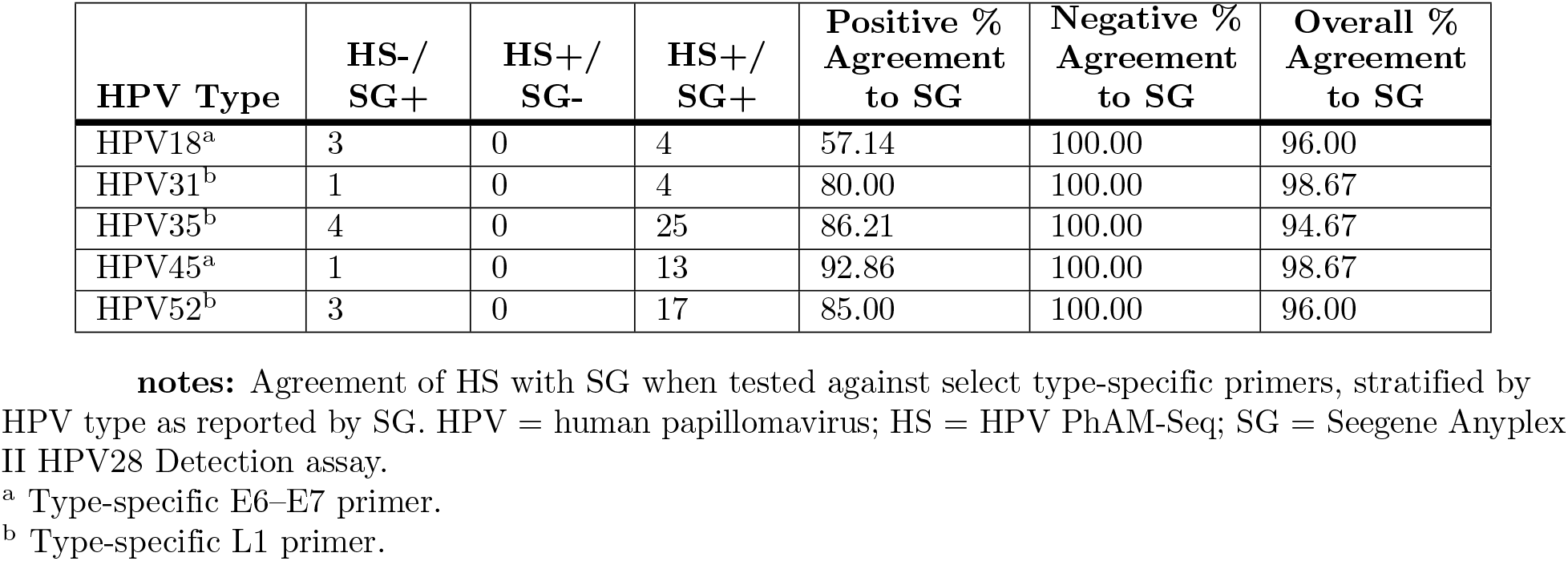
Agreement of HS with SG when tested against select type-specific primers, stratified by HPV type as reported by SG.

| HPV Type | HS-/SG+ | HS+/SG- | HS+/SG+ | Positive % Agreement to SG | Negative % Agreement to SG | Overall % Agreement to SG |
| --- | --- | --- | --- | --- | --- | --- |
| HPV18<sup>a</sup> | 3 | 0 | 4 | 57.14 | 100.00 | 96.00 |
| HPV31<sup>b</sup> | 1 | 0 | 4 | 80.00 | 100.00 | 98.67 |
| HPV35<sup>b</sup> | 4 | 0 | 25 | 86.21 | 100.00 | 94.67 |
| HPV45<sup>a</sup> | 1 | 0 | 13 | 92.86 | 100.00 | 98.67 |
| HPV52<sup>b</sup> | 3 | 0 | 17 | 85.00 | 100.00 | 96.00 |

<sup>a</sup> Type-specific E6–E7 primer.
<sup>b</sup> Type-specific L1 primer.

All 14 SG-typed HPV45 samples were tested with custom primers targeting the HPV45 E6–7 locus. HPV45 was detected in all but one sample, resulting in a positive percent agreement of 92.9%, compared to 14.3% with MY11/09 primers.

Seven of the 14 SG-typed HPV18 samples were tested using type-specific primers also targeting the E6–7 locus. The overall positive percent agreement for this subset was 57.1% (compared to a positive percent agreement of 35.7% with MY11/09 primers).

Of the 27 SG-typed HPV52 mono-infected samples in the blinded cohort, 20 were evaluated using custom type-specific primers targeting the L1 gene locus. Seventeen samples (85%) were identified as HPV52-positive, compared to 55.6% using MY11/09 primers. Of the 3 samples in which HPV52 DNA was not detected by HPV PhAM-Seq, 2 were also determined to be HPV52-negative with MY11/09 primers in the blinded validation cohort, while the remaining sample was typed as HPV58.

### HPV PhAM-Seq has the capacity to detect novel variants of common HPV types

Commercial PCR-based assays can distinguish among HPV types but do not resolve intra-type genetic variation. In contrast, HPV PhAM-Seq provides genotypic information at single-nucleotide resolution, enabling identification of distinct variants within the same HPV type. To identify variants, our analysis pipeline determines the most abundant paired read (Read 1/Read 2) sequence combination for each sample assigned at least one HPV type. All unique read-pair combinations for a given HPV genotype observed across samples are compiled and assigned a subtype identifier. Each sample is then annotated with the subtype ID corresponding to its dominant HPV type. Using this approach, we identified two distinct subtypes of HPV16 and HPV31 with the degenerate MY primer set, and two to three distinct subtypes each of HPV35, HPV45, and HPV52 using E6–E7 type-specific primers. Importantly, although different individuals harbored different subtypes, the same subtype was consistently detected across all samples collected from a given individual (Fig 4). Notably, one read assigned to HPV45 subtype 2 had no perfect matches in the nr database, suggesting this subtype represents a previously unreported variant of HPV45.

**Figure 4.**
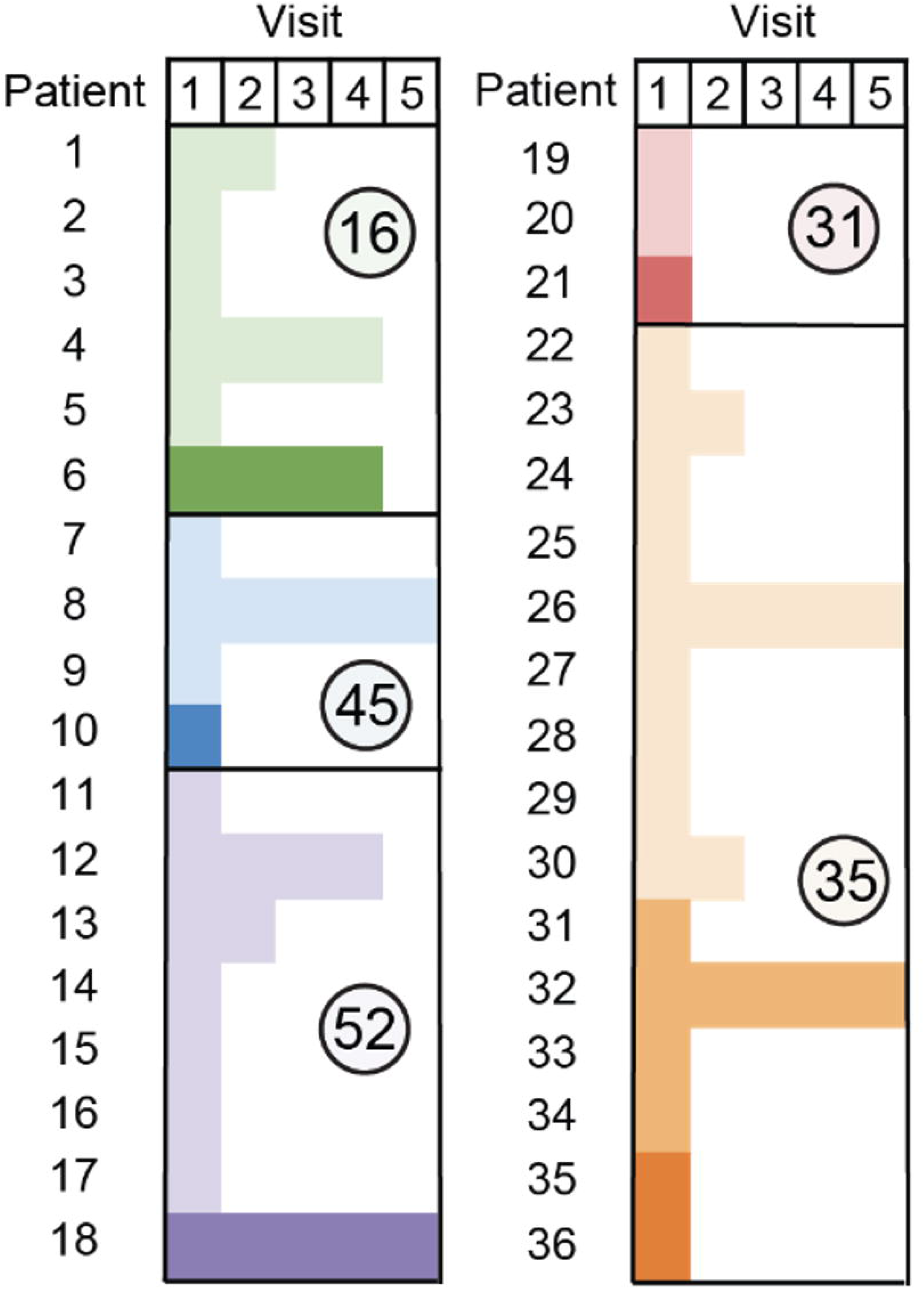
The distribution of HPV16/31/35/45/52 subtypes detected across multiple samples from the same individual. Within each HPV type, different shades represent unique subtypes.

### Non-normalization of samples prior to pooling results in similar HPV genotyping performance compared to the original protocol

To streamline assay workflow, we assessed the effect of omitting sample normalization between enrichment steps on HPV PhAM-Seq performance. In the non-normalized workflow, equal volumes of samples with DNA concentrations greater than 0.1ng/ul after the first enrichment were pooled together, regardless of concentration. Eliminating this normalization step reduced hands-on processing time by an estimated 90 seconds per sample or pool (including dilution calculations, preparation and pipetting time) and eliminated the need for post-enriched quantification and size determination. Pooling samples directly after the first PCR reduced labor and consumable requirements associated with quantification, sample dilution, and normalization (i.e., pipette tips, reservoirs, strip tubes). We compared assay performance against SG for both normalized and non-normalized samples. Positive-percent agreement with SG was lower in the non-normalized workflow for most high-risk HPV types (HPV16/31/45/52/58), with reductions ranging from 7.2% – 37.5% relative to the normalized workflow (Supplementary Table 4).

### HPV PhAM-Seq can perform HPV genotyping for <20 USD per sample, even at smaller scales

HPV-PhAM-Seq can be performed at relatively low per-sample cost across a range of sequencing scales and Illumina platforms (Table 5). At the smallest scale, the estimated, combined cost of DNA extraction, library preparation and sequencing for 400 samples on an iSeq 100 is $10.90 per sample, whereas use of the higher-throughput NovaSeq X platform with greater multiplexing capacity reduced the estimated per-sample cost to $8.56 USD (including $0.02 USD per sample for sequencing). For the 170 samples in our blinded validation cohort, the estimated library construction and sequencing cost was $9.65 per sample, plus an estimated additional $7.36 per sample for extraction with automation.

**Table 5.**
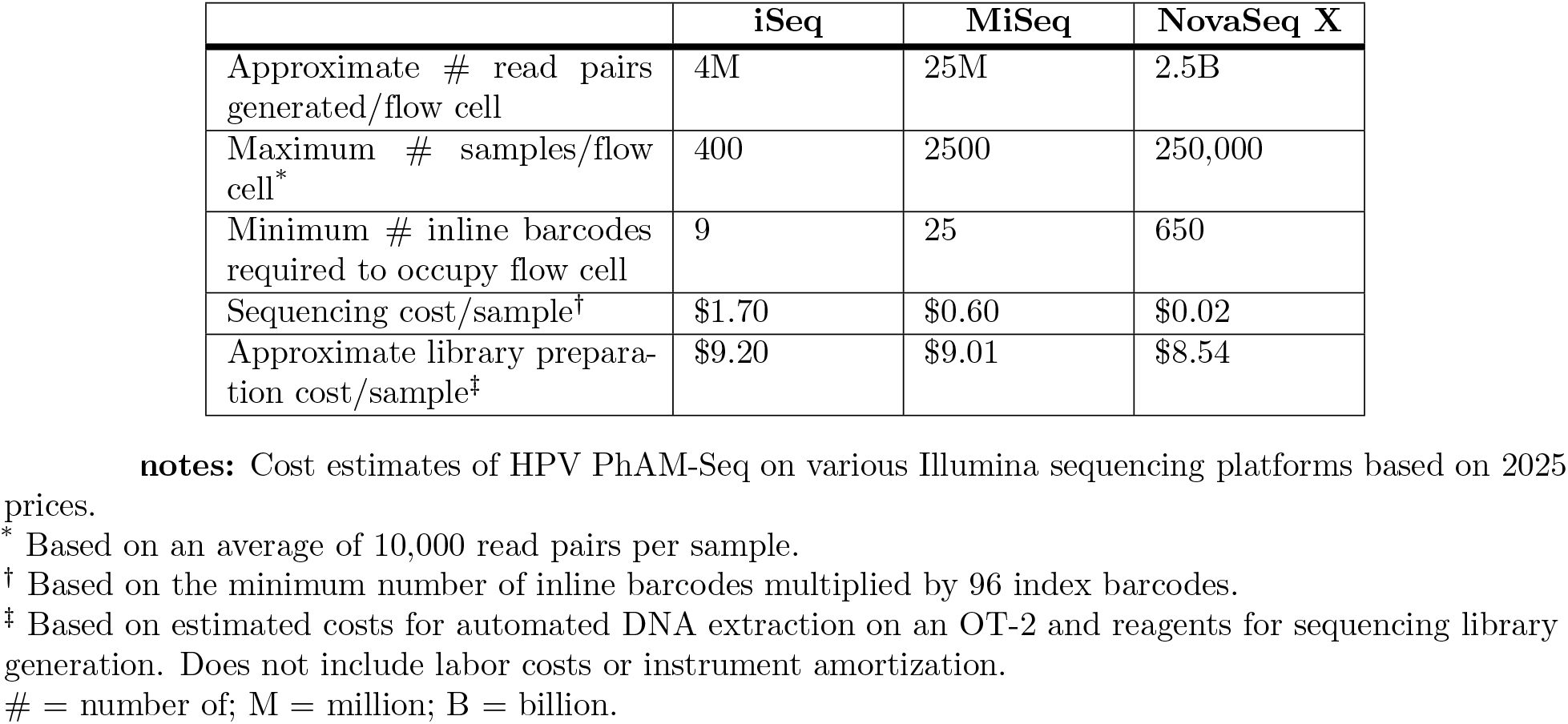
Cost estimates of HPV PhAM-Seq on various Illumina sequencing platforms based on 2025 prices.

|  | iSeq | MiSeq | NovaSeq X |
| --- | --- | --- | --- |
| Approximate # read pairs generated/flow cell | 4M | 25M | 2.5B |
| Maximum # samples/flow cell* | 400 | 2500 | 250,000 |
| Minimum # inline barcodes required to occupy flow cell | 9 | 25 | 650 |
| Sequencing cost/sample† | \$1.70 | \$0.60 | \$0.02 |
| Approximate library preparation cost/sample‡ | \$9.20 | \$9.01 | \$8.54 |

\* Based on an average of 10,000 read pairs per sample. † Based on the minimum number of inline barcodes multiplied by 96 index barcodes. ‡ Based on estimated costs for automated DNA extraction on an OT-2 and reagents for sequencing library generation. Does not include labor costs or instrument amortization.

## Discussion

Despite robust evidence supporting the effectiveness of primary hrHPV screening to reduce cervical cancer rates, PCR-based hrHPV testing still fails to meet the scale and cost parameters required for its sustainable implementation in parts of the globe with the highest cervical cancer burden. Meanwhile, reductions in the cost of at-scale sequencing has facilitated an opportunity for next generation sequencing-based HPV genotyping of populations. We developed PhAM-Seq as a flexible, scalable, and cost-effective amplicon sequencing platform for HPV genotyping. This platform uses amplicons tagged with unique combinations of phased inline barcodes and Illumina indexes, enabling hundreds to thousands of samples to be pooled for library construction and sequencing. In this study, we validated HPV PhAM-Seq (HS) against the Seegene Anyplex II HPV28 system (SG) on a cohort of 170 human cervical samples, observing *>*80% overall agreement for the 8 most common hr genotypes associated with invasive cervical cancer. HPV PhAM-Seq demonstrated the highest positive percent agreement with SG for types HPV16, HPV33, and HPV58 using the MY11/09 degenerate primer set. Performing HPV PhAM-Seq with type-specific primers instead of MY11/09 improved capture of most of the remaining types (i.e., HPV18/35/45/52), with a range in positive percent agreement across all hr types tested that was more narrow and greater in value (i.e., 57.2–92.86% with type-specific primers compared to 3.3–81.3% with MY primers), paving the way for further optimization efforts to improve hr type-specific sensitivity.

The application of NGS technology for infectious disease diagnostics continues to grow, and its role in HPV genotyping for both cervical cancer screening and characterizing type distribution in populations is increasingly being recognized [28, 41, 42]. Despite this trend, PCR-based assays continue to dominate the spectrum of HPV diagnostic testing in current use, despite the reality that 50% of HPV tests on the global market lack a single peer-reviewed publication and *>*75% lack reassuring data on its performance on key testing characteristics [24]. While internationally recognized quality standards and best-practice guidelines are essential to facilitate the adoption of next-generation sequencing (NGS) [41], several additional barriers may limit its broader implementation. These include the technical and analytical complexity of existing NGS platforms, as well as the associated costs [42]. Few publications have reported cost estimates for NGS HPV diagnostics and of those reported, most are at least $50 USD per sample (Table 1). Specific platforms that may overcome these cost barriers [26] remain hindered by long processing times and difficulty to replicate. Additionally, many publications describing these NGS platforms offer relatively sparse detail on operationalizing the assay, including types of primers used [26] or their methodology for sequencing data analysis [26, 27].

PhAM-Seq aims to address each of these barriers by providing a detailed standard operating procedure (SOP) for laboratory workflows; an accessible Google Workspace–based system for primer design, sample tracking, and metadata management; and an end-to-end analysis pipeline that integrates sample and process metadata with sequencing data, enabling rapid assignment of HPV types and subtypes as well as comparison with results from previous typing assays. Lowering these barriers to adoption is particularly important for LMICs that shoulder most of the global burden of cervical cancer. It took one day to perform DNA extraction for 182 samples, with 4.7 hours of hands-on time, while HPV PhAM-Seq took 4 days with 17 hours of hands-on time. Although this turnaround time is longer than that of PCR-based assays, it is comparable to other HPV NGS platforms [26, 27, 43]. Importantly, this timeframe aligns with the standard care cascade for cervical cancer screening and follow-up, where further evaluation and treatment of hrHPV-associated cervical disease typically does not happen on the day of hrHPV diagnosis [44]. Further, this timeline may help mitigate overtreatment associated with more recently-proposed point-of-care hrHPV testing and same-day treatment approaches [45].

Additionally, the HPV PhAM-Seq platform can be tuned for greater sensitivity or specificity by modulating thresholds of one or more parameters, depending on the use case (e.g., typing in a restricted-versus abundant-resource setting, where different levels of permissiveness for false-negatives exist). Primer sets can be substituted or added to the PhAM-Seq system without changing its workflow. Furthermore, the PhAM-Seq platform, by design, can be adapted to type pathogens other than HPV by modifying the primer target.

The cost estimate of running HPV PhAM-Seq on our blinded validation was *<*$20 USD per sample, lower than the $50 USD pricepoint of many other NGS HPV diagnostics, even accounting for labor and instrument costs. Still, this price overestimates the projected per-sample cost when the platform is scaled to maximal capacity on an iSeq 100 system or higher throughout Illumina sequencing platforms. When comparing the cost of HPV PhAM-Seq to non-NGS HPV diagnostics currently in wider use in LMICs, this platform still performs competitively, approaching the estimated price point of $14 necessary for HPV-based cervical cancer screening strategies to be more cost-effective than cytology [46]. Furthermore, the price estimate at scale–even when accounting for additional costs such as sample collection, instrument maintenance, labor and supply chain–remains comparable to or lower than the negotiated per-test costs of various commercial assays currently used in LMICs [47] while providing the added value of sequencing-based genotyping results.

Further, like other NGS diagnostic platforms, HPV PhAM-Seq offers the additional advantage of sequence-level typing data. Unlike PCR-based assays, whose targets are predetermined based on the primers used, HPV PhAM-Seq can theoretically detect any HPV genotype and variations of genotypes, as long as sequence loci are identified with primer-binding regions that are sufficiently conserved across genotypes and intervening regions that are sufficiently divergent across genotypes. For example, HPV PhAM-Seq using MY primers detected either HPV71 or HPV70 in the majority of the SG-identified HPV35 samples that were successfully typed, and it detected either HPV72 or HPV70 in majority of typed, SG-identified HPV45 samples. While the use of type-specific primers with HPV PhAM-Seq also indicated the presence of HPV35 or HPV45 DNA in these samples, our typing results suggest that HPV70/71/72 may commonly co-infect with HPV35/45. The association of HPV70 with other high-risk types has been previously described [48, 49], but less is known about HPV71/72. Notably, HPV71/72 are low risk types not present on the SG panel; HPV70 is on the SG panel and is classified as low-risk, while emerging evidence suggests probable carcinogenicity [50]. Further, among types that were positive by HPV PhAM-Seq and negative by SG, HPV16, HPV33, and HPV58 were more commonly detected relative to other types. Studies have shown reduced agreement between Seegene Anyplex HPV28 compared to other reference assays for HPV16, HPV33, and HPV58, suggesting increased intra-type sequence variation for these genotypes that may compromise SG performance [51]. The nucleotide-level data provided by HPV PhAM-Seq also allows for identification of novel variants of established genotypes, particularly for HPV16, HPV31, HPV35, HPV45, and HPV52, adding to the collective knowledge of intra-type variation and allowing for potential determination of persistent versus new infection [41, 42]. Therefore, HPV PhAM-Seq could serve as an epidemiologic tool for HPV typing of populations, including vaccine impact monitoring, in addition to primary cervical cancer screening.

HPV PhAM-Seq harbors limitations. Most importantly, the MY11/09 primer set used in our blinded validation failed to bind and amplify all 8 RG types equally. The limitation in the targeting capacity of MY has been previously described and these primers, which were first described in 1989 [52], have since been redesigned to improve its sensitivity for a wider range of HPV genotypes. Examples include the HMB01 [53] and PGMY [54] primer sets, the latter of which has been demonstrated to detect specifically HPV35/45/52 with greater frequency compared to standard MY11/09 (Gravitt et al. Improved amp of genital HPV. JCM. PMID: 10618116). In previous experiments (data not included), we tested human cervical samples from this cohort with PGMY primers and were unable to replicate this improved detection of these types compared to MY11/09. This finding may have been influenced by various factors, including using a different DNA polymerase than what is used in the current iteration of the platform (i.e., Platinum II Taq Hot-Start DNA Polymerase). We also observed off-target amplification and poor detection of certain types when combining MY11/09 with type-specific primers targeting the same L1 region as MY (data not included), which may be due to biased amplification in specimens with HPV co-infections [55] or failure to detect types due to L1 gene disruption from HPV integration into human chromosomal DNA [56].

Future iterations of the platform can address these issues through multiple approaches. One approach is to trial combining MY11/09 with E6–7 type-specific primers, which has demonstrated improved HPV detection in PCR-based assays by targeting different regions of the HPV genome [57, 58]. Alternatively, to minimize any potential challenges with off-target amplification or primer dimerization from increasing primer multiplexity, an alternative approach would be to conduct a “first pass” iteration of HPV PhAM-Seq on all samples using MY11/09 primers, and then, for all hrHPV-negative samples, re-run HPV PhAM-Seq using type-specific primer(s). The particular type-specific primers could be customized based on a combination of the hr genotypes most likely missed by consensus primers and those that are most prevalent in the testing population. This two-step approach may incur additional costs, but assuming an hr HPV positivity rate of approximately 0.3 among women in Sub-Saharan Africa per earlier population studies [59] and the inherently greater capacity for scale with this technology, per-sample costs may yet remain competitive with other HPV testing assays in LMICs. Finally, it is also possible to utilize newer bioinformatic tools [60] to design entirely new consensus, degenerative, or multiplexed primer sets that can target a wider range of genotypes than MY alone.

Additional limitations include the fact that, as with any validation study where an assay is tested against a reference standard, the true diagnostic value of HPV PhAM-Seq may be undermined by limitations in SG, whereby the HPV genes targeted by its primers are not specified [61]. Further, this platform was not validated on self-collected samples, which is increasingly used in HPV testing [42]. Lastly, when applied to cervical cancer screening, this platform would not function as a point-of-care assay, and therefore would not allow for same-day testing and treatment.

## Conclusion

In conclusion, to address the need for massively increased capacity for population-level HPV screening, we developed an economical, scalable and readily implementable NGS amplicon sequencing platform for HPV genotyping with *>*80% overall agreement to a validated, commercial assay for the 8 most common genotypes associated with invasive cervical cancer with a single degenerative primer set targeting the L1 gene. Sensitivity of this platform for HPV35 and HPV45 was increased by 83% and 78%, respectively, with the utilization of type-specific primers. The NGS nature of HPV PhAM-Seq offers a number of advantages, including 1) detecting HPV genotypes beyond the pre-determined targets of PCR-based tests, 2) identifying novel variants, 3) distinguishing new versus persistent infections by detecting nucleotide-level variation within HPV genotypes, and 4) accounting for potential human chromosomal integration events resulting in L1 gene loss by also working with primers targeting the E6–7 gene loci. In a moment where the major hurdles facing wider adoption of hrHPV-based cervical cancer screening are cost, access, and the technical complexities with implementation, HPV PhAM-Seq offers a uniquely cost-effective, transparent, and readily implementable NGS HPV genotyping tool, primed for a range of global public health applications from cervical cancer screening to population characterization and vaccine impact monitoring.

## Data Availability

All data produced in the present study are available upon reasonable request to the authors

The HPV PhAM-Seq analytical pipeline developed for this study is publicly available through GitHub and includes all Python and Unix shell scripts, command-line workflows, reference files, documentation, workbooks for sample tracking and primer design, and associated resources required to reproduce the computational analyses described in this manuscript. The version of the repository corresponding to this preprint has been permanently archived in Zenodo (DOI: 10.5281/zenodo.21631215) to ensure long-term accessibility and reproducibility.

https://github.com/dipeshsolanky91-debug/HPV-PhAM-Seq-Toolbox

## Acknowledgments

The authors express their gratitude to Sofia Fatouros, along with members of the Microbial Omics Core and Bhattacharyya Lab at the Broad Institute for their support and cooperation during this study. The authors also thank the research support staff at the University of Washington as well as the study participants and investigators of MTN-020/ASPIRE for provision of samples used for assay validation.

## Supporting information

**S1 Table.**
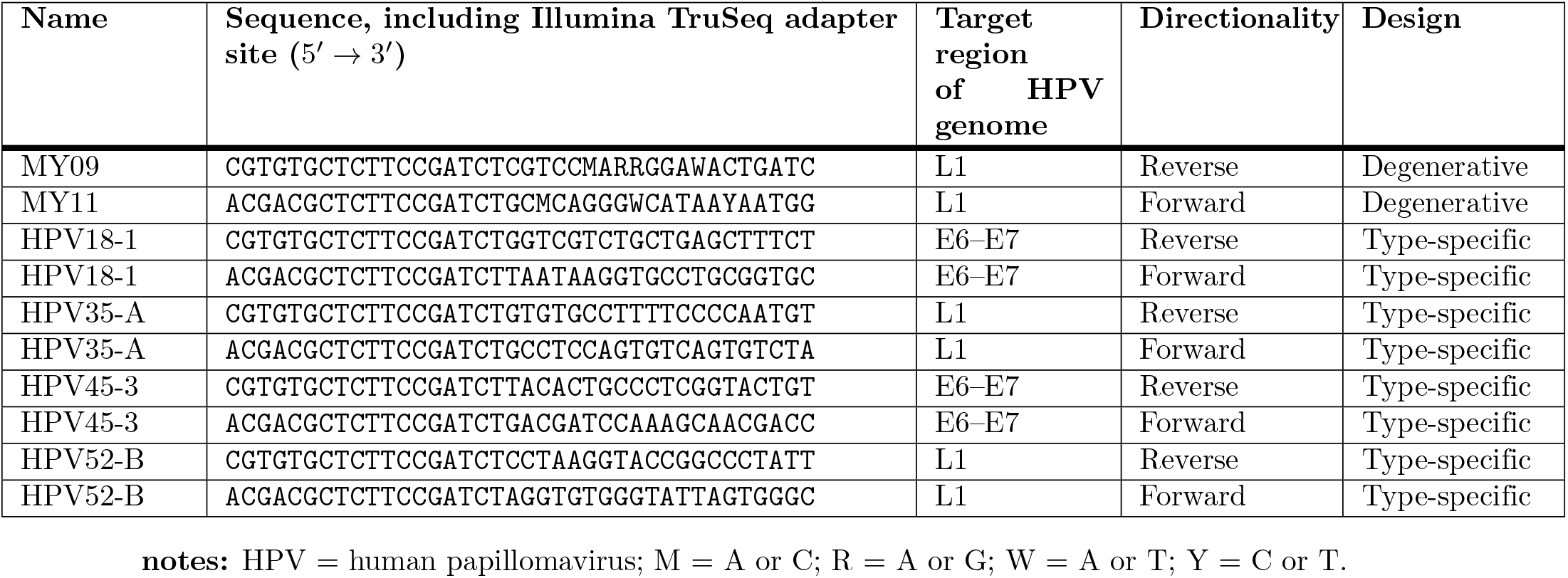
HPV-Specific primer sequences used in HPV PhAM-Seq library preparation.

**S2 Table.**
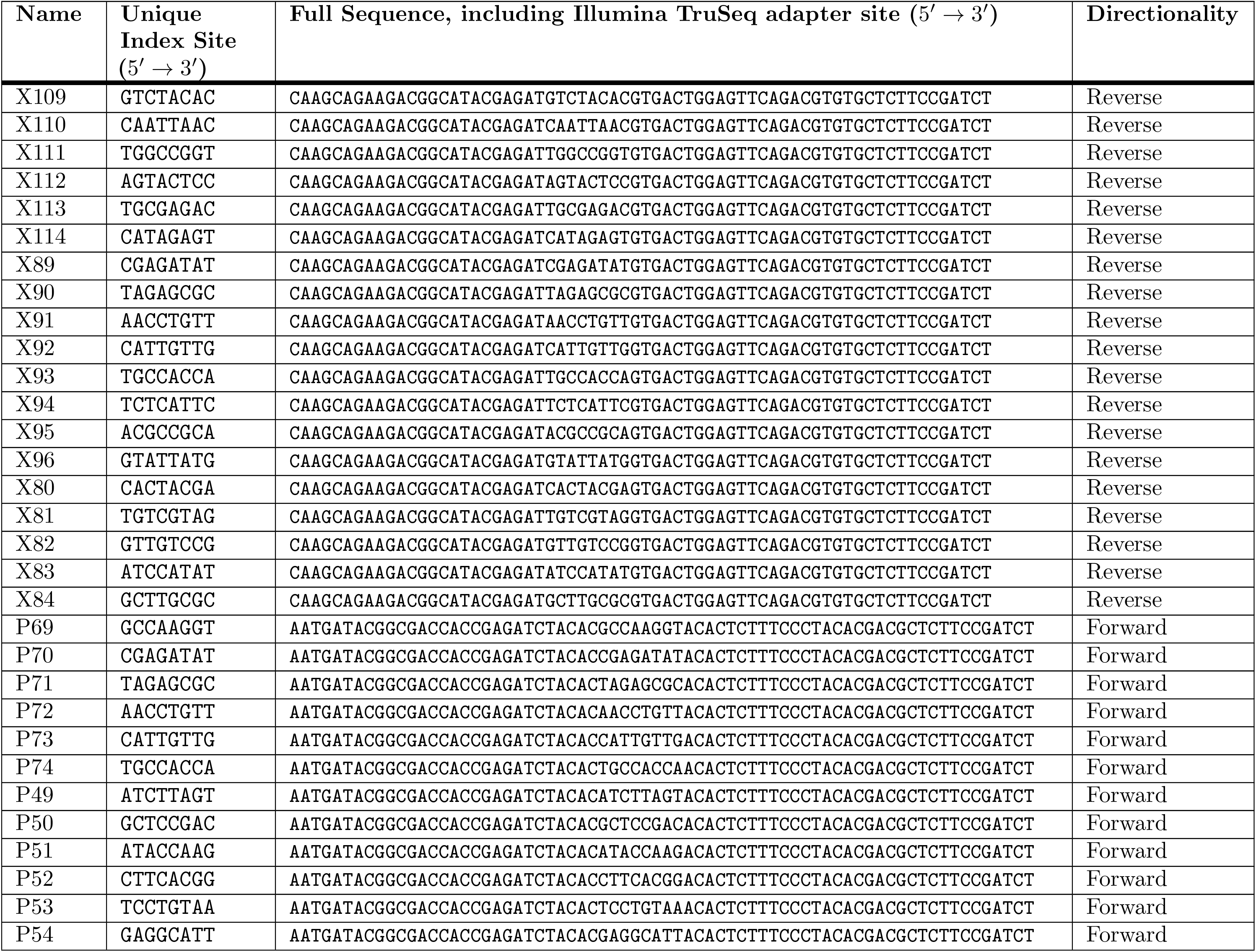
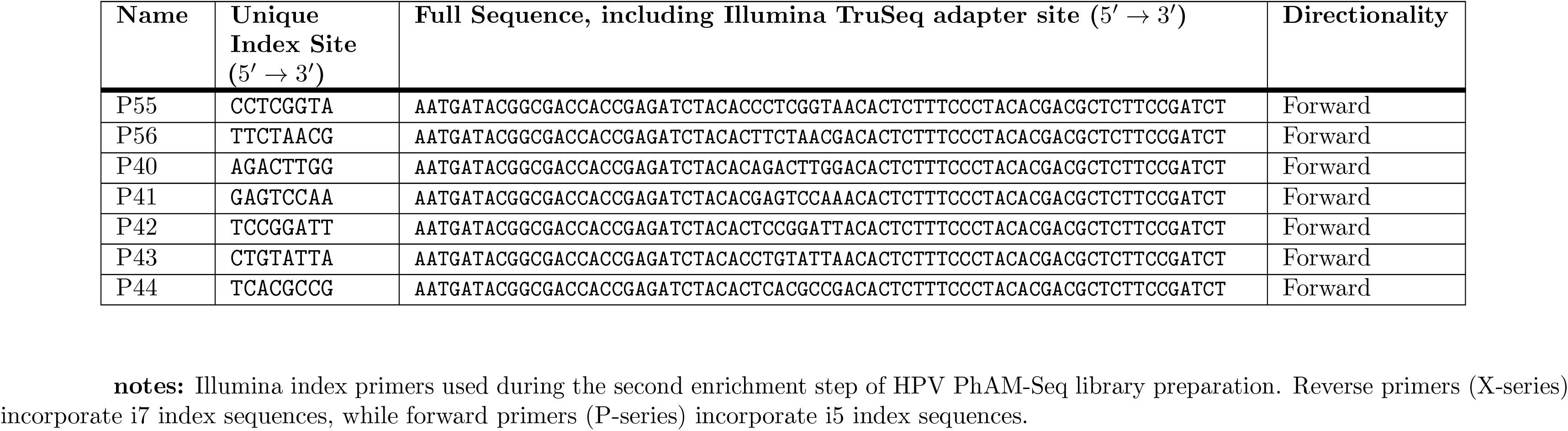
Illumina index primer sequences used in HPV PhAM-Seq library preparation.

## S1 Appendix. Generation of HPV L1, E6, and E7 sequence database

For each accession and gene in the curated HPV reference database, only the longest CDS was retained to remove redundancy, and HPV types were assigned to each gene using BLASTn against a curated HPV reference database based on the highest bitscore hit subject to gene-specific minimum alignment lengths (L1 *geq*1000 bp, E6 *geq*350 bp, E7 *geq*200 bp). Assigned types were embedded into FASTA headers using the format TYPE:GENE:ACCESSION. Identical nucleotide sequences were collapsed to produce non-redundant gene-specific FASTAs, which were then concatenated to generate the final combined HPV L1E6E7 reference database for downstream alignment and typing analyses. This database contained 3936 unique sequences (2763, 791, and 382 sequences for L1, E6, and E7, respectively) for a total of 195 different HPV types. The database along with all scripts, workbooks, reference files and documentation related to the pipeline is available as described in the Data Availability Statement.

### Sequencing data processing and HPV genotyping: supplemental information

Paired-end FASTQ files were processed using a custom primer-aware parser (HPV bc parse) to detect primer sequences and determine staggered offsets. Reads lacking detectable primer sequences were excluded from downstream demultiplexing. Barcode-based demultiplexing was performed using a paired-end demultiplexer (hpv bc demux pe) using sequence-specific assignment (SEQ SPEC=Y), requiring exact matches of both stagger length and barcode sequence on R1 and R2. Matched read pairs were written to barcode-specific FASTA files, capped at a predefined maximum read count per barcode to normalize coverage.

Reads were aligned to our curated HPV L1E6E7 database of non-redundant L1, E6, and E7 gene sequences (see above) using BWA. For reads with multiple alignments, the primary alignment was selected based on highest mapping quality and lowest mismatch count. As previously described, per-sample HPV types were summarized by counting reads assigned to each type and a sample was associated with an HPV type(s) if the sample reached or exceeded thresholds specified on the command line. For each sample, all the identified HPV types were reported, along with the percentage of HPV reads assigned to each type. Concordance with SG was classified as follows: POS M1/POS M2/POS M3 (SG-classified type same as first/second/third highest abundance type), POS MM (SG-classified type different than first/second/third highest abundance type), FNEG (SG-classified with an HPV type, no HPV detected), and TNEG (no HPV detected by SG or by HPV PhAM-Seq). Among the samples assayed, no identified HPV types in the fourth highest abundance or lower matched to the HPV type detected by SG. The entire script for the pipeline and all command lines are available as described in the Data Availability Statement.

### Validation of HPV PhAM-Seq on a blinded cohort (additional information)

Samples (n=156) with concentrations of 0.1 ng/uL or more after the first enrichment were pooled and used as input for the second enrichment. Of the 14 samples not sequenced due to insufficient concentration following the first enrichment, 7 were samples that SG identified as HPV35, followed by HPV-negative (n=3), HPV18 (n=2), HPV16 (n=1) and HPV33 (n=1). The higher proportion of HPV35 not amplified was unsurprising as this genotype is typically not well amplified by MY11/09 primers [62]. The average quantification of samples after the first enrichment was 9.78 ng/uL (n=156, median 6.91, range 0.15–40.08). Prior to the second enrichment, samples were grouped into 19 pools comprising 7–9 samples per pool. The sequencing run yielded 1.77 GB of data. A total of 89.2% of reads achieved quality scores of *geq*Q30. The sequencing run for the non-normalized assay generated 1.59 GB of data, with 89.8% of reads with quality scores of *geq*Q30.

### Advantage of phased, inline barcodes for highly multiplexed sequencing

The use of short, phased (variable-length), inline barcodes in PhAM-Seq provides several advantages. First, inline barcoding substantially increases multiplexing capacity. For example, we designed 6 unique inline barcodes (IBCs) of 0–2 nt length such that each differs in length and/or by a Hamming distance of at least two from all others. This enables 36 (6 × 6) unique forward-reverse IBC combinations using 12 PhAM-Seq primers. When each pool is further amplified with a standard set of Illumina primers containing 96 unique combinations of index sequences, thousands of samples can be multiplexed within a single sequencing lane. Further, if the maximum staggered IBC length is increased to 3 nucleotides, the number of unique IBCs goes up to 22, and the number of unique forward-reverse IBC combinations increases to 484 allowing for the multiplexing of tens of thousands of samples. Second, long 5’ primer overhangs elevate the risk of inter- and intra-primer interactions and off-target hybridization, potentially reducing amplification efficiency and fidelity [63, 64]. By minimizing the length of inline barcodes and of the universal Illumina sequence incorporated into the initial primers, PhAM-Seq mitigates these risks. Finally, amplicon sequencing frequently suffers from low base diversity, which can compromise sequencing quality. Although diversity can be artificially increased by spiking in unrelated libraries (e.g., PhiX), it is challenging to add sufficient diversity without sacrificing substantial sequencing capacity. By shifting the amplicon sequence relative to the sequencing initiation site (Figure 1C), PhAM-Seq IBCs increase base diversity across sequencing cycles even when the underlying amplicon sequences are highly uniform [65].

**S1 Fig. Discordant cases between HS and SG** Columns represent the type called by SG in the upper grid and those identified by HS in the lower grid. Numbers in the upper grid do not represent a single sample as samples are counted more than once if multiple types were detected by HS. HPV=human papillomavirus; HS=HPV PhAM-Seq; SG=Seegene Anyplex II HPV28 Detection assay.

**S4 Table.** Exact McNemar tests assessing discordance between HPV PhAM-Seq and SG with versus without sample normalization.

| HPV Type | Sensitivity<br>McNemar $\chi^2$ | Sensitivity<br>*p*-value (Exact) | Specificity<br>McNemar $\chi^2$ | Specificity<br>*p*-value (Exact) |
| --- | --- | --- | --- | --- |
| HPV16 | 3 | 0.25 | 3 | 0.25 |
| HPV18 | 0 | 1 | 0 | 1 |
| HPV31 | 3 | 0.25 | 1 | 1 |
| HPV33 | 0 | 1 | 0 | 1 |
| HPV35 | 0 | 1 | 0 | 1 |
| HPV45 | 1 | 1 | 0 | 1 |
| HPV52 | 8 | 0.007812 | 1 | 1 |
| HPV58 | 6 | 0.03125 | 0.2 | 1 |
| HPV-negative | 0 | 1 | 24 | 0 |

