## Supplementary figures and images for "Phased amplicon multiplex sequencing for cost-effective detection of high-risk human papillomavirus from cervical samples"

### Supplemental Figure 1

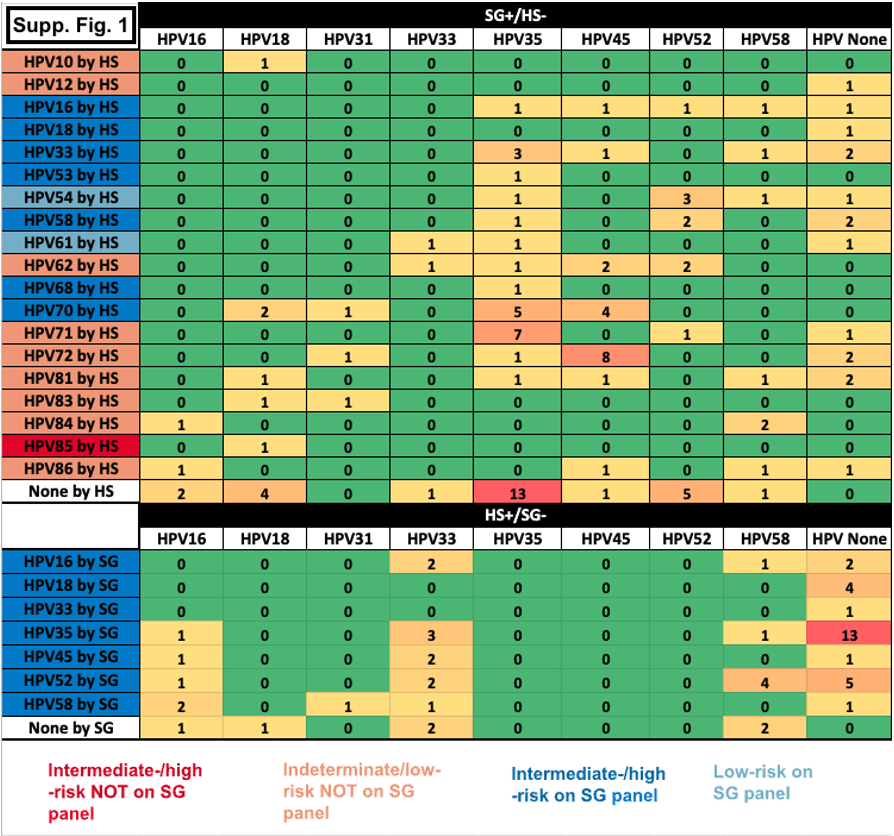
